# Changes in ischemic stroke presentations and associated workflow during the first wave of the COVID-19 pandemic: A population study in Alberta, Canada

**DOI:** 10.1101/2021.10.04.21264529

**Authors:** Aravind Ganesh, Jillian M. Stang, Finlay A. McAlister, Oleksandr Shlakhter, Jessalyn K. Holodinsky, Balraj Mann, Michael D. Hill, Eric E. Smith

## Abstract

**Background:** Pandemics may promote hospital avoidance among patients with emergencies, and added precautions may exacerbate treatment delays. There is a paucity of population-based data on these phenomena for stroke. We examined the effect of the COVID-19 pandemic on the presentation and treatment of ischemic stroke in an entire population.

**Methods:** We used linked provincial administrative data and data from the Quality Improvement and Clinical Research Alberta Stroke Program – a registry capturing stroke-related data on the entire population of Alberta(4.3 million)– to identify all patients presenting with stroke in the pre-pandemic(1-January-2016 to 27-February-2020, n=19,531) and pandemic(28-February-2020 to 30-August-2020, n=2,255) periods. We examined changes in thrombolysis and endovascular therapy(EVT) rates, workflow, and in-hospital outcomes.

**Results:** Hospitalizations/presentations for ischemic stroke dropped (weekly adjusted-incidence-rate-ratio[aIRR]:0.48, 95%CI:0.46-0.50, adjusted for age, sex, comorbidities, pre-admission care needs), as did population-level incidence of thrombolysis(aIRR:0.49,0.44-0.56) or EVT(aIRR:0.59,0.49-0.69). However, the proportions of presenting patients receiving acute therapies did not decline (e.g. thrombolysis:11.7% pre-pandemic vs 13.1% during-pandemic, aOR:1.02,0.75-1.38). Onset-to-door times were prolonged; EVT recipients experienced longer door-to-reperfusion times (median door-to-reperfusion:110-minutes, IQR:77-156 pre-pandemic vs 132.5-minutes, 99-179 during-pandemic; adjusted-coefficient:18.7-minutes, 95%CI:1.45-36.0). Hospitalizations were shorter but stroke severity and in-hospital mortality did not differ.

**Interpretation:** The first COVID-19 wave was associated with a halving of presentations and acute therapy utilization for ischemic stroke at a population level, and greater pre-hospital and in-hospital treatment delays. Our data can inform public health messaging and stroke care in current and future waves. Messaging should encourage attendance for emergencies and stroke systems should re-examine “code stroke” protocols to mitigate inefficiencies.

## INTRODUCTION

In response to the Coronavirus Disease 2019 (COVID-19) pandemic, affected countries implemented varying degrees of social-distancing measures to decrease viral transmission. An unintended consequence of social-distancing messaging could be hospital avoidance by patients with emergencies like acute coronary syndrome (ACS) or stroke, as previously observed in Taiwan and Hong Kong during outbreaks in 2003 and 2009 respectively.^1,2^ Some of the groups most worried about contracting COVID-19 are at high cardiovascular risk, like older people or those with heart disease who were specifically warned of being at higher risk of severe COVID-19 in some public health messaging.^3^ Social distancing may also result in loss of services and support networks for seniors or patients with disabilities, potentially impairing their ability to seek medical assistance.^4^ Furthermore, pandemics generate additional challenges of managing personal protective equipment.(PPE) and cleaning protocols,^5^ and additional information bottlenecks, all of which could result in pre-hospital and in-hospital workflow delays for emergencies like stroke.^6^

There are preliminary reports of a decline in patients presenting to hospital with acute stroke or ACS.^7,8^ A World Stroke Organization survey of members in multiple countries indicated a sharp reduction in acute stroke admissions by up to 50-80% in the first few weeks of the pandemic.^9^ An international cross-sectional study reported a global decline in stroke hospitalizations.^10^ These reports are especially troubling because recent viral or respiratory infections are known to double the risk of stroke and ACS,^11^ suggesting we should be seeing more, not fewer, of these presentations. There is also some indication that those patients who do present are doing so later than usual, perhaps because they are waiting until their condition becomes more severe or intolerable.^12,13^ Two small studies from Hong Kong reported delays in presentation for patients with stroke and ACS seen during the COVID-19 pandemic compared to the year before.^14,15^

Verifying and quantifying the effect of the pandemic on stroke presentations and workflow can help us better tailor our public health messaging to continue emphasizing the time-critical nature of emergencies like stroke. Such data may also help us optimize our pandemic stroke workflow protocols, or at least adjust our expectations for what can be accomplished under such exceptionally challenging circumstances. We undertook a population study of changes in hospitalizations, treatment rates, presentation delays, and workflow delays for ischemic stroke during the first wave of the COVID-19 pandemic in the province of Alberta, Canada.

## METHODS

We used linked provincial administrative data and data from the Quality Improvement and Clinical Research (QuICR) Alberta Stroke Program, a registry capturing stroke-related data on the entire population of Alberta.^16^ Data on all patients hospitalized with ischemic stroke in Alberta during our Pre-Pandemic and Pandemic periods of interest (defined below) were obtained from the Discharge Abstract Database (DAD) maintained by the provincial health authority (Alberta Health Services Analytics group via the Alberta Strategy for Patient Oriented Research Support.Unit, AbSPORU) which contains demographic and clinical information on hospital discharges, including comorbidities and need for continuing care like assisted living or nursing home care.^17^ In addition, we obtained data on all emergency department visits for minor stroke (i.e. ischemic stroke discharged home directly from emergency) in Alberta from the National Ambulatory Care Reporting System (NACRS). DAD and NACRS data are reliable in reporting ischemic stroke and vascular risk factors,^18^ with International Classification of Diseases (ICD)-10 codes for stroke (**Supplement**) having accuracies of 92-97%.^19^

In addition, we extracted data on use of thrombolysis and endovascular therapy (EVT) through the QuICR program, which includes two comprehensive stroke centres (the sole providers of EVT) and 15 urban and remote hospitals designated as Alberta’s primary stroke centres, which can provide thrombolysis. No other centres provide hyperacute stroke care. The QuICR registry, released in January 2016, collects key data for all patients receiving thrombolysis and/or EVT (details at https://cumming.ucalgary.ca/research/quicr/home). It is managed by stroke co-ordinators at each of the 17 stroke centres. Quality control measures are taken quarterly; the data are comprehensively validated against electronic medical records. We abstracted patient demographics; stroke severity (National Institutes of.Health Stroke Scale score [NIHSS]); pre-hospital workflow (times for stroke onset, call to emergency medical services, ambulance dispatch, paramedics-on-scene, hospital arrival); and in-hospital workflow or treatment-related metrics (CT, thrombolysis, EVT groin puncture and reperfusion times). These data were linked using the patients’ provincial healthcare number.

The study was approved by the University of Calgary Conjoint Health Research Ethics Board (REB20-0769). No informed consent was required.

### Statistical Analyses

Based on our sample size calculations **(Supplement)**, we needed at least 38 treated patients in the Pandemic period to achieve 80% power for identifying a 25% drop in treatment volume or times. By 30-August-2020, we had already accrued 408 patients in QuICR during the pandemic, so we proceeded with our analyses.

Alberta’s first COVID-19 case was identified on 28-February-2020.^20^ In March, the Government of Alberta declared a state of emergency under the Public Health Act and imposed measures limiting gatherings to 15 people, prohibiting and/or limiting attendance at public and private facilities. Daycares were closed, educational institutions switched to online learning, and companies switched to work-from-home arrangements whenever possible. All “close-contact” health and personal care services were closed, and scheduled non-urgent and elective surgeries were postponed.^21^ Therefore, the “Pandemic period” for this study was defined as 28-February-2020 to 30-August-2020 (inclusive) whereas the “Pre-Pandemic period” was defined as 1-January-2016 to 27-February-2020.

For the primary analyses, we compared the rate of patients presenting to emergency departments with stroke and the rate of thrombolysis/EVT for stroke (as cases over time) in the Pre-Pandemic and Pandemic periods using Poisson regression, adjusted for age, sex, continuing care needs, and (for thrombolysis/EVT) NIHSS and onset-to-door time. The decision to pursue this pre- and post-analysis – as opposed to various potential methods like interrupted time series, other segmented regression modelling, or restricting the pre-pandemic period to certain months – was made based on published provincial stroke data, which have shown that there is no seasonality to stroke admissions in Alberta.^22^ For the incidence rate ratio calculation in the Poisson regressions, we defined time epochs as weeks (e.g. stroke hospitalizations/presentations per week for Pandemic vs Pre-Pandemic periods), and then verified our findings using monthly and quarterly time epochs as well.

For secondary analyses of pre-hospital factors, we compared the proportions of all stroke patients who received thrombolysis/EVT in the Pandemic vs Pre-Pandemic periods using (a) Fisher’s exact and (b) logistic regression adjusted for age, NIHSS, and onset-to-door time. As EVT utilization was expected to increase over the study period given expanding awareness and indications, particularly with the publication of late-window trials in 2018,^23^ we performed a sensitivity analysis restricting the Pre-Pandemic comparison period to 1-January-2018 onwards. We used similar logistic regressions for in-hospital mortality. We also compared pre- and in-hospital workflow times and hospital length-of-stay using (a) Wilcoxon rank sum and (b) generalized linear models (GLM) adjusted for age, sex, pre-admission continuing care needs, comorbidities, and NIHSS. We compared the NIHSS on admission in the Pandemic vs Pre-Pandemic periods using Wilcoxon rank sum and GLM adjusted for age, sex, comorbidities, and continuing care needs.

Analyses of incidence and onset-to-call/door times were stratified by age (≥65-years vs <65-years) and pre-admission continuing care needs (any vs none) after testing for heterogeneity of effects, as we were particularly interested if there were differences among these groups. All analyses were performed with STATA/MP 16.1. Statistical significance was defined as p<0.05.

## RESULTS

We analyzed 19,531 patients with ischemic stroke pre-pandemic versus 2,255 during the pandemic (**Table 1**). Patients who presented during the pandemic were less likely to have atrial fibrillation, hypertension, and chronic kidney disease and were less likely to have ≥2 comorbidities, even after adjusting for age, sex, and continuing care needs pre-hospitalization (adjusted odds-ratio [aOR]:0.85, 95%CI:0.77-0.94).

**Table 1.** Key demographic and clinical characteristics of patients presenting or hospitalized with ischemic stroke in the Pre-COVID-19-Pandemic (1-January-2016 to 27-February-2020) and Pandemic periods (28-February-2020 to 30-August-2020) in Alberta, Canada.

| Characteristic | Pre-Pandemic period | Pandemic period |
| --- | --- | --- |
| **Entire study population** | N=19,531 | N=2,255 |
| Hospitalized, N (%) | 15,344 (78.6) | 1,805 (80.0) |
| Minor stroke (non-hospitalized), N (%) | 4,187 (21.4) | 450 (20.0) |
| Age, median (IQR) | 72 (60-82) | 71 (61-82) |
| Sex – female, N (%) | 9,101 (46.6) | 1,059 (47.0) |
| **Continuing care needs, N (%)** |  |  |
| Any continuing care | 2,645 (13.5) | 304 (13.5) |
| Nursing home care | 485 (2.5) | 43 (1.9) |
| **Comorbidities, N (%)** |  |  |
| Atrial fibrillation | 2,632 (17.7) | 238 (13.7) |
| Coronary Artery Disease | 275 (1.9) | 32 (1.9) |
| Chronic Kidney Disease | 404 (2.7) | 28 (1.6) |
| Diabetes Mellitus | 4,298 (28.9) | 508 (29.3) |
| Heart Failure | 634 (4.3) | 60 (3.5) |
| Hypertension | 9,141 (61.6) | 1,000 (57.7) |
| Any of above | 11,027 (56.5) | 1,238 (54.9) |
| ≥2 of above (multi-morbidity) | 5,184 (26.5) | 531 (23.6) |
| **Acute treatment, N (%)** |  |  |
| Thrombolysis | 2,277 (11.7) | 295 (13.1) |
| Endovascular therapy (EVT) | 1,081 (5.5) | 159 (7.1) |
| Hospital length-of-stay, median days (IQR) | 7 (3-19) | 6 (3-13) |
| In-hospital mortality (%) – excluding non-hospitalized minor stroke | 1,614 (10.6) | 176 (9.9) |

Hospitalizations/presentations for ischemic stroke dropped in the Pandemic period, whether using weekly, monthly, or quarterly epochs (**Figure 1, Table 2**). There was no interaction by age (p-interaction:0.48), pre-admission continuing care needs (p-interaction:0.63), or minor stroke diagnoses (vs major stroke, p-interaction:0.41). There were similar declines in the incidence of thrombolysis and EVT, compared to the somewhat stable overall incidence of thrombolysis pre-pandemic (**Figure 2A**) and the previously rising utilization of EVT **(Figure 2B, Table 2)**.

**Table 2.**
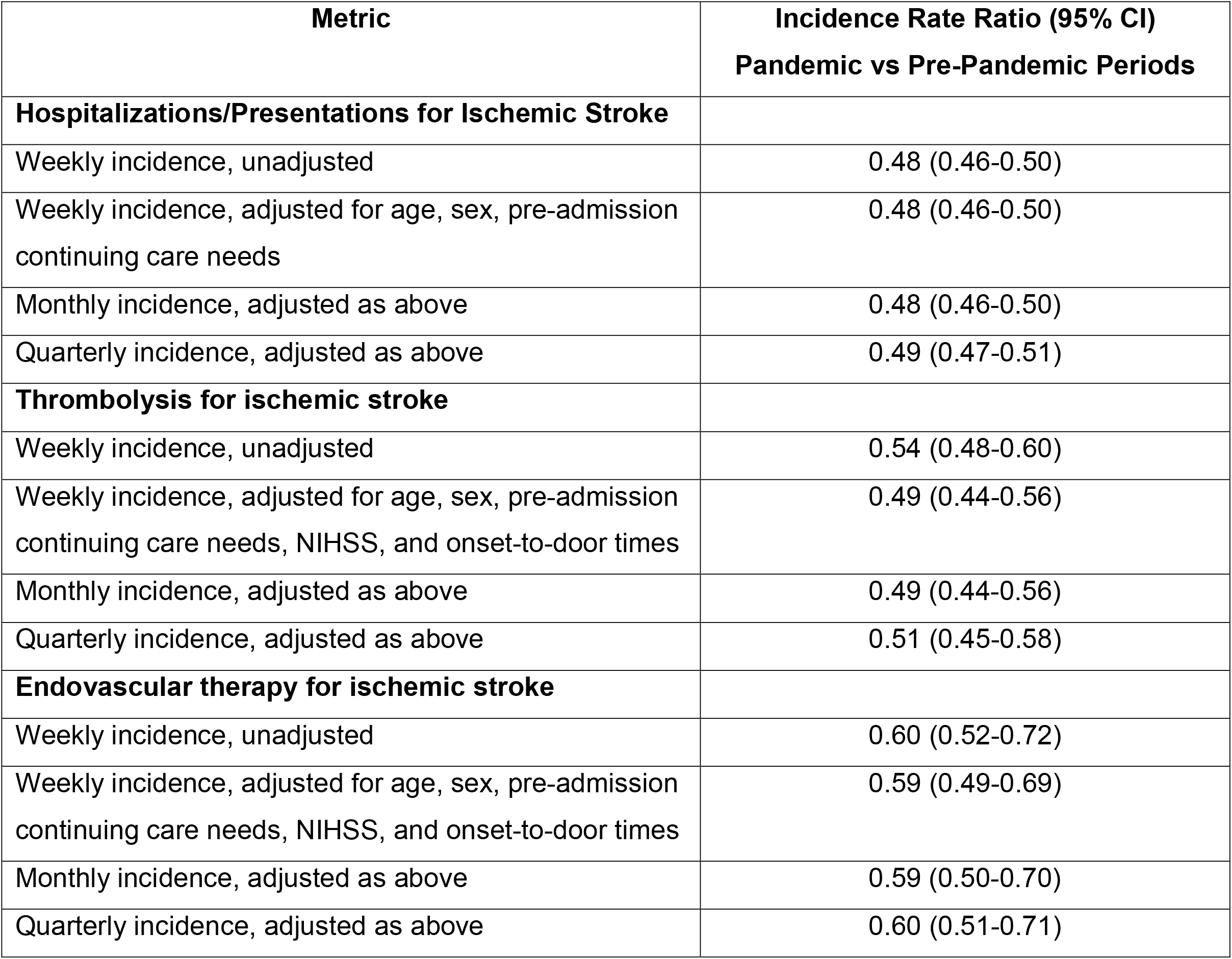
Change in the incidence of hospitalizations/presentations and acute therapies for ischemic stroke in the Pandemic (28-February-2020 to 30-August-2020) versus Pre-Pandemic (1-January-2016 to 27-February-2020) Periods in Alberta, Canada.

| **Metric** | **Incidence Rate Ratio (95% CI)**<br>**Pandemic vs Pre-Pandemic Periods** |
| --- | --- |
| **Hospitalizations/Presentations for Ischemic Stroke** |  |
| Weekly incidence, unadjusted | 0.48 (0.46-0.50) |
| Weekly incidence, adjusted for age, sex, pre-admission continuing care needs | 0.48 (0.46-0.50) |
| Monthly incidence, adjusted as above | 0.48 (0.46-0.50) |
| Quarterly incidence, adjusted as above | 0.49 (0.47-0.51) |
| **Thrombolysis for ischemic stroke** |  |
| Weekly incidence, unadjusted | 0.54 (0.48-0.60) |
| Weekly incidence, adjusted for age, sex, pre-admission continuing care needs, NIHSS, and onset-to-door times | 0.49 (0.44-0.56) |
| Monthly incidence, adjusted as above | 0.49 (0.44-0.56) |
| Quarterly incidence, adjusted as above | 0.51 (0.45-0.58) |
| **Endovascular therapy for ischemic stroke** |  |
| Weekly incidence, unadjusted | 0.60 (0.52-0.72) |
| Weekly incidence, adjusted for age, sex, pre-admission continuing care needs, NIHSS, and onset-to-door times | 0.59 (0.49-0.69) |
| Monthly incidence, adjusted as above | 0.59 (0.50-0.70) |
| Quarterly incidence, adjusted as above | 0.60 (0.51-0.71) |

**Figure 1.**
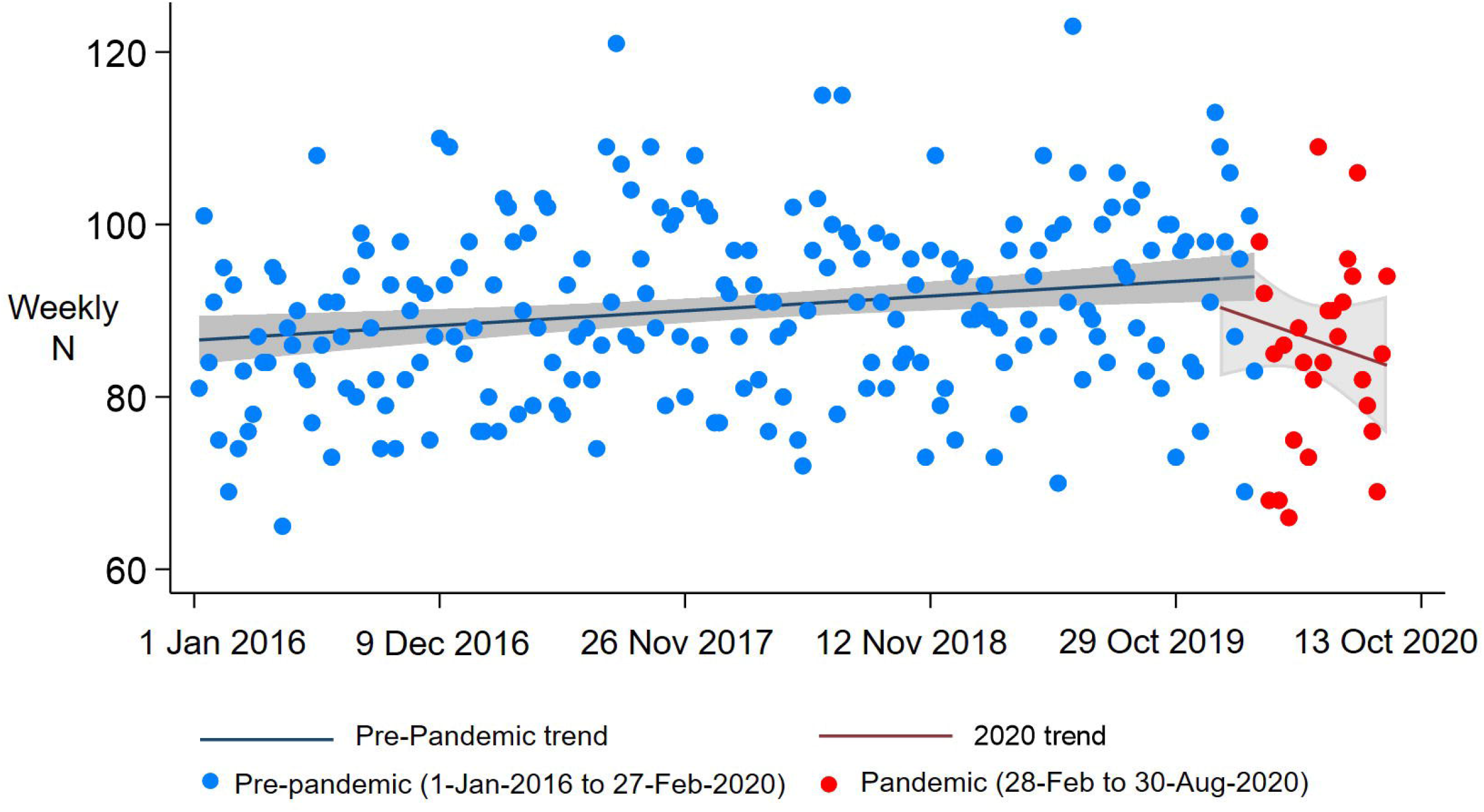
Weekly incidence of hospitalizations/presentations for ischemic stroke in Alberta, Canada in the Pre-Pandemic and Pandemic periods. The Pre-Pandemic period was defined as 1-January-2016 to 27-February-2020 whereas the Pandemic period extended from 28-February-2020 to 30-August-2020. The x-axis is marked off in 50-week increments. Trendlines from linear regression are shown for each period of interest, with shaded 95% confidence intervals.

**Figure 2.**
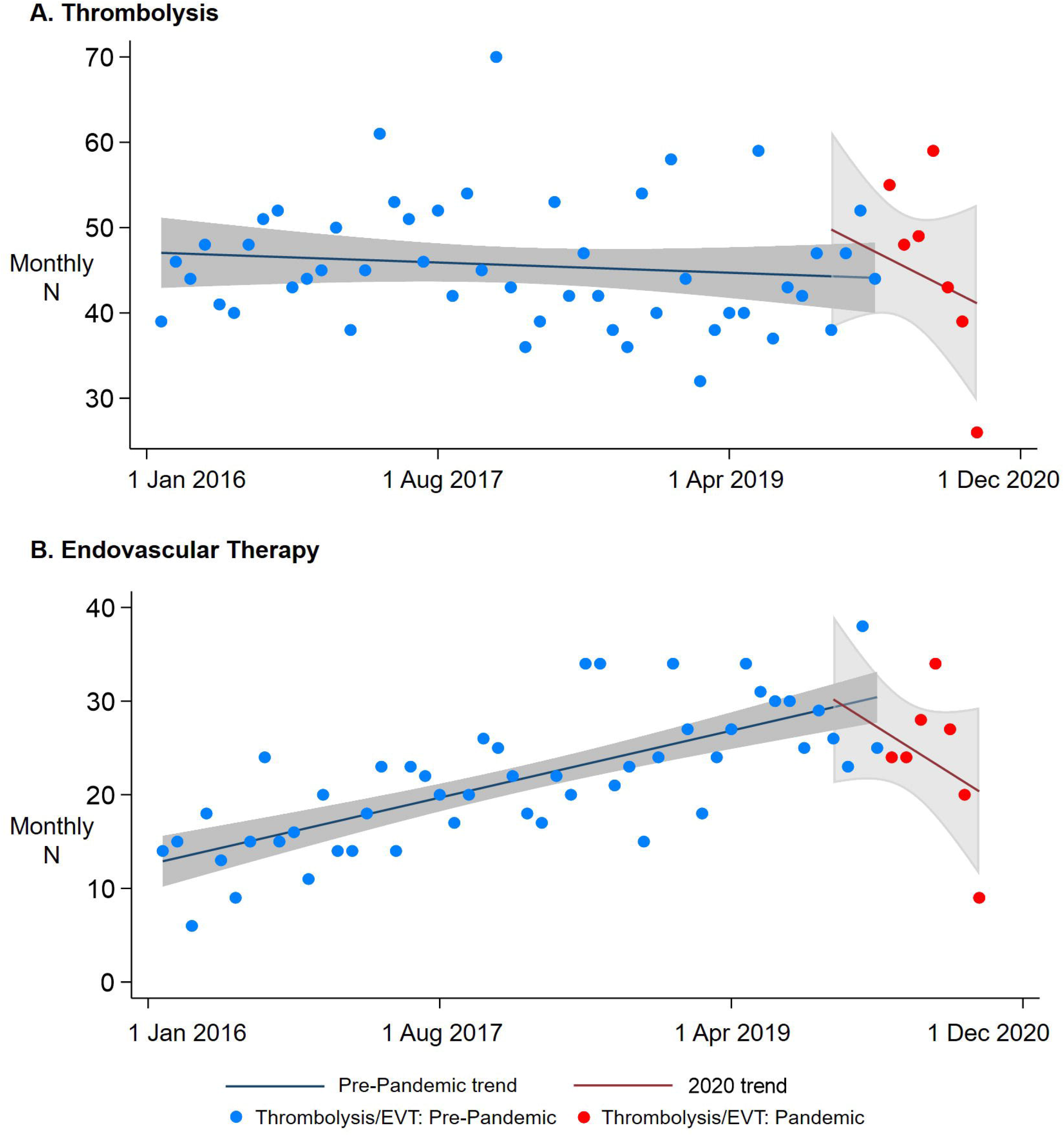
Monthly incidence of (A) thrombolysis and (B) endovascular therapy (EVT) for ischemic stroke in Alberta, Canada in the Pre-Pandemic and Pandemic periods. The Pre-Pandemic period was defined as 1-January-2016 to 27-February-2020 whereas the Pandemic period extended from 28-February-2020 to 30-August-2020. The x-axis is marked off in 20-month increments. Trendlines from linear regression are shown for each period of interest, with shaded 95% confidence intervals.

However, the proportions of patients presenting with an ischemic stroke receiving thrombolysis and EVT did not decline during the first wave (**Table 1**, odds-ratio adjusted for age/NIHSS[aOR]:0.83,0.63-1.08; EVT aOR:1.41,1.11-1.80). Whereas the proportion of patients receiving EVT generally increased over the study period, upon restricting the pre-COVID-19 period to 2018 onwards (EVT given to 730[7.0%] of 10,506 patients), no difference was seen (aOR:1.16,0.90-1.48). Among those receiving EVT, thrombolysis was given to 505(46.7%) of 1,081 patients pre-pandemic vs 77(48.4%) of 159 patients during the pandemic (aOR:1.13,0.80-1.59). Similar results were seen on further adjusting for onset-to-door times, except that the apparent higher odds of EVT in the pandemic period were attenuated (thrombolysis aOR:1.02,0.75-1.38, EVT aOR:1.28,1.00-1.65).

Onset-to-door times could be calculated for all 3,048 treated patients with out-of-hospital ischemic stroke, and were prolonged during the pandemic (**Table 3**). This effect was not modified by age or continuing care needs. Among 2,532(98.6%) thrombolysis recipients and 1,146(92.4%) EVT recipients, there were no significant differences in door-to-CT or door-to-needle times, but CT-to-groin-puncture, door-to-groin-puncture, groin-puncture-to-reperfusion, and door-to-reperfusion times for EVT were prolonged post-COVID-19 (**Table 3**). Differences in door/CT-to-groin-puncture times were attenuated on adjusted GLM, but delays in groin-puncture-to-reperfusion and door-to-reperfusion remained significant (**Table 3**).

**Table 3.**
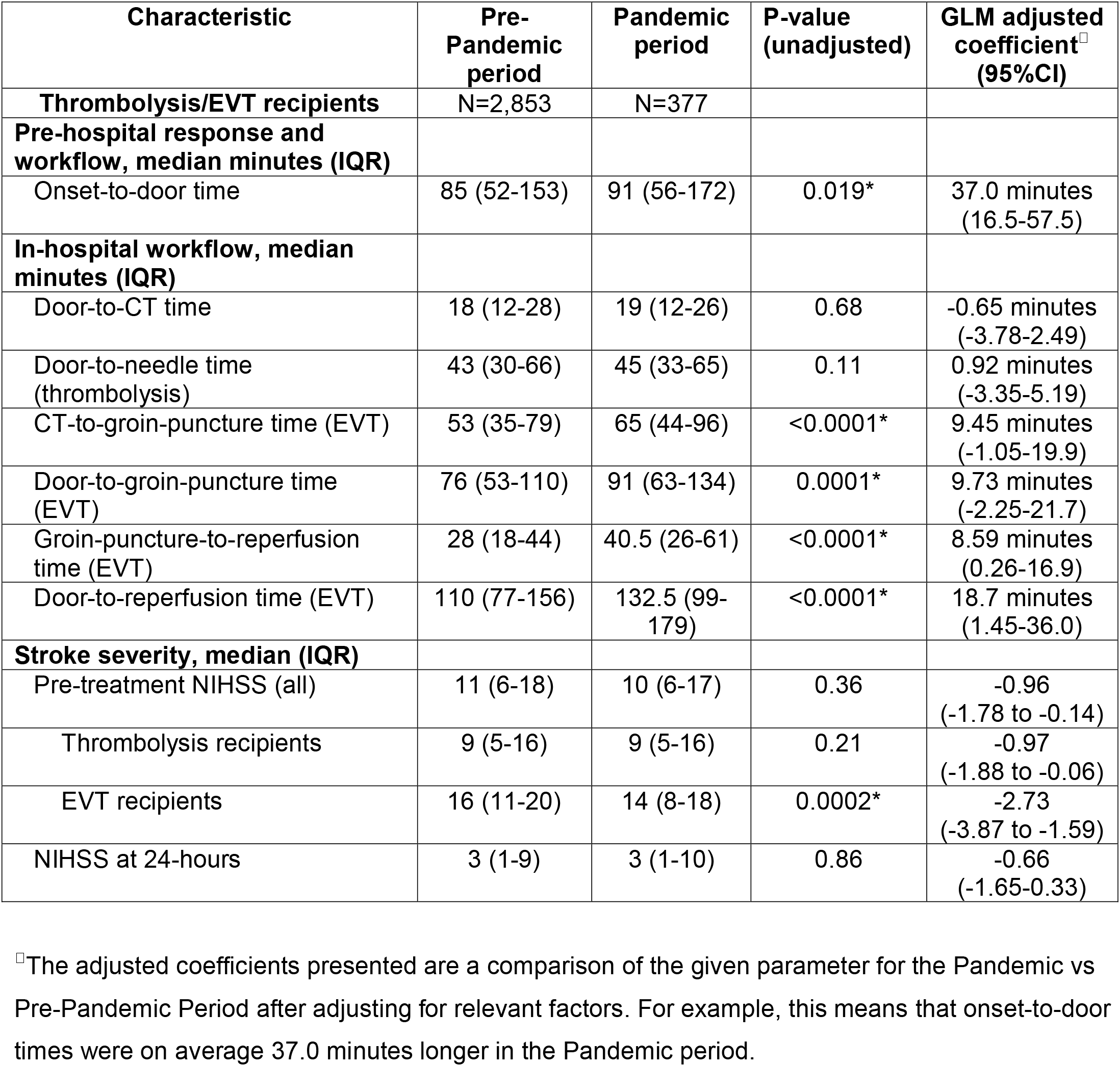
Pre-hospital and in-hospital factors and stroke severity of patients with ischemic stroke who received acute stroke therapies in the Pre-COVID-19-Pandemic (1-January-2016 to 27-February-2020) and Pandemic periods (28-February-2020 to 30-August-2020) in Alberta, Canada. Significant differences (p<0.05) for hypotheses testing are indicated with an asterisk (*). Generalized Linear Models (GLMs) were adjusted for age, sex, pre-admission continuing care needs, comorbidities, and (for all except NIHSS analyses) the baseline NIHSS.

Among patients treated with thrombolysis or EVT, there were no differences in NIHSS scores at baseline on unadjusted analyses (**Table 3**). For modelling analyses, we imputed the baseline NIHSS for 33 patients using the 24-hour NIHSS score. There was a slightly lower NIHSS among treated patients during the pandemic after adjusting for age/sex/continuing-care needs (**Table 3**), modified by receipt of EVT (p-interaction for EVT recipients=0.018), such that the NIHSS was about 2 points lower among EVT recipients in the pandemic vs pre-pandemic periods (adjusted-coefficient:-2.16, −3.20 to − 1.11). This reflected secular trends in offering therapies to “milder” patients, as findings were attenuated after adjusting for time epochs (adjusted-coefficient: −0.72, −1.62-0.19).

Lengths-of-stay were shorter during the pandemic (**Table 1**, p<0.0001), including on adjusting for age/sex/continuing care/comorbidities/NIHSS (adjusted-coefficient: −6.61 days, −10.6 to −2.63). There was no difference in in-hospital mortality (**Table 1**), including after adjusting for age, sex, continuing care, comorbidities (aOR:0.97, 0.81-1.15, p=0.68) and when restricting to treated patients, adjusting additionally for NIHSS (aOR:0.94, 0.62-1.44, p=0.78).

## DISCUSSION

Using the population of an entire Canadian province, we found that the first wave of the COVID-19 pandemic was associated with a marked decline in hospital presentations for ischemic stroke and in the absolute utilization of acute stroke therapies including thrombolysis and EVT. There were greater delays in hospital presentation and in both pre-hospital and in-hospital workflow. We have validated concerns raised by different groups from survey data about the pandemic’s indirect effects on medical emergencies.^24,25^ Declines on the order of 40% or greater in ACS and stroke admissions were reported from surveys of Spanish centres.^26,27^ A cross-sectional study from the United States found that ACS hospitalizations decreased by about 19 cases/week, with a rise in risk-adjusted mortality.^13^ Preliminary reports on stroke care included a 25% relative drop in stroke admissions and 14% drop in thrombolysis among Italian stroke units in March 2020 versus March 2019.^8^ We have now shown that the pandemic was associated with a similar magnitude of decline in presentations for ischemic stroke and use of acute therapies, even after adjusting for relevant variables and including/excluding minor stroke. It is unlikely that the drop reflects a true decline in stroke occurrence, and we suspect that it reflects pandemic-related hospital avoidance, as also indicated by a recent population-based study showing fewer abdominal and gynecological emergency department visits in Ontario during the pandemic.^28^ Importantly, the diminished utilization of thrombolysis/EVT reflected the decline in stroke presentations rather than any therapeutic inertia: the proportion of presenting patients receiving thrombolysis was unchanged and that receiving EVT increased over the study period, reflecting pre-pandemic trends.

Pre-hospital delays in onset-to-door times confirm that patients who presented with acute stroke during the first wave tended to present later than usual. Observed delays are in keeping with prior small studies reporting delayed stroke and ACS presentations.^14,15^ Despite the additional delays, however, we did not find any increase in stroke severity (per NIHSS) in the pandemic period. An unconfirmed hypothesis is that.higher rates of out-of-hospital stroke deaths might lie at the root of.the decrease in average NIHSS scores of those presenting to hospital, in keeping with reported increases in out-of-hospital cardiac arrests.^29,30^ Nevertheless, our data should motivate healthcare systems to ensure that emergency treatment indications like stroke are not drowned out by pandemic-related public health messaging. Striking the right balance is challenging, but as a general principle, it may be prudent for public health agencies to avoid blanket stay-at-home statements that may inadvertently promote hospital avoidance. Instead, strong statements promoting social distancing or other such measures could be combined with messages reminding and encouraging people to still seek immediate medical attention for emergencies like stroke.

Workflow delays at both the pre-hospital and in-hospital stages (particularly for EVT) reflect the procedural changes necessary for pandemic care (use of PPE for all staff, enhanced cleaning protocols for imaging equipment and procedure rooms, etc). Preliminary data on such delays have come from different parts of the world for ACS^13,31^ and acute stroke,^32^ including a single-center Canadian study which reported longer onset-to-door, door-to-needle, and door-to-reperfusion times.^33^ Whereas some delays may be inevitable under such challenging circumstances, our population data should serve as a call to action for stroke systems to closely re-examine their pandemic protocols to mitigate workflow inefficiencies wherever possible.

Whereas a key strength of our study is that the data arise from an entire population with a coordinated and robust stroke system of care, a few limitations merit discussion. First, we cannot know to what extent observed delays or declines in presentations were related to difficulties calling for help or accessing medical services, versus pure hospital avoidance. Second, because the cohort was defined by ICD administrative data, strokes may have been misclassified in some cases; however, our data sources and case definitions have been validated in prior audits.^18^ Third, data on pre-hospital and in-hospital workflow were only available for patients who received acute stroke therapies, likely resulting in underestimation of delays, since patients presenting much later would be less likely to receive therapies like thrombolysis. Whereas both hospitals and paramedics adopted additional infection prevention precautions, we do not have specific data on why delays occurred in different settings.

Fourth, we do not have longer-term outcome data on most patients in the pandemic era, so we could not assess downstream effects of observed delays; however, post-stroke outcomes are highly dependent on time-to-treatment.^34^ Fifth, our dataset did not contain the COVID-19 status of patients, so we do not know whether patients were managed differently based on their infectious state; however, all patients were managed as potentially infectious in the first 24-hours pending RT-PCR results. Furthermore, rates of COVID-19 in Alberta were very low during the first wave, with about 14,000 cases or 0.35% of the population affected;^20^ thus, this is almost purely a study of the effects of public health measures, patient avoidance, and hospital infection prevention measures, in the absence of major strains on emergency medical services or hospitals from large numbers of infected patients.

In conclusion, the first wave of the COVID-19 pandemic in Alberta was associated with a halving of presentations for ischemic stroke and use of acute therapies at a population level, and greater pre-hospital and in-hospital workflow delays, but in those patients who did present, the provision of thrombolysis/EVT was similar to pre-pandemic levels. Given our findings, there seems little doubt that the cerebrovascular implications of the COVID-19 pandemic will extend beyond just direct infection-related damage in survivors. Public health messaging in future waves should encourage hospital attendance for acute conditions like stroke rather than blanket stay-at-home statements.

## Supporting information

STROBE checklist

## Data Availability

The data that support the findings of this study are available from the corresponding author upon reasonable request.

## Supplement

Supplemental Methods. STROBE and RECORD Checklists separately attached.

## CONTRIBUTORS

AG conceived and co-designed the study, secured funding, performed statistical analysis and interpretation, and wrote and revised the manuscript. JMS collected data for the study and contributed to the analysis. FAM co-designed the study, provided resources for data collection, and contributed to both data interpretation and manuscript revision. OS, JKH, and BM helped design the study and provided critical revision of the manuscript. MDH and EES co-designed the study, provided study supervision and funding, interpreted data, and revised the manuscript.

