## Supplemental Methods for "Changes in ischemic stroke presentations and associated workflow during the first wave of the COVID-19 pandemic: A population study in Alberta, Canada"

**Codes used for identifying cases**

ICD-10 diagnostic codes were used to identify all patients who presented to hospitals or emergency departments in Alberta during the study period with a final diagnosis of ischemic stroke for their episode of care. An episode of care refers to all contiguous inpatient hospitalizations. These case identification codes are based on the Heart and Stroke Foundation’s Canadian Best Practice Recommendations (2016).

| ICD-10-CA Code | Description |
| --- | --- |
| Ischemic Stroke | |
| G08 | Intracranial and Intraspinal Phlebitis and Thrombophlebitis |
| H341 | Central Retinal Artery Occlusion |
| I630 | Cerebral Infarction Due To Thrombosis Of Precerebral Arteries |
| I631 | Cerebral Infarction Due To Embolism Of Precerebral Arteries |
| I632 | Cerebral Infarction Due To Unspecified Occlusion Or Stenosis Of Precerebral Arteries |
| I633 | Cerebral Infarction Due To Thrombosis Of Cerebral Arteries |
| I634 | Cerebral Infarction Due To Embolism Of Cerebral Arteries |
| I635 | Cerebral Infarction Due To Unspecified Occlusion Or Stenosis Of Cerebral Arteries |
| I636 | Cerebral Infarction Due to Cerebral Venous Thrombosis, Nonpyogenic |
| I638 | Other Cerebral Infarction |
| I639 | Cerebral Infarction, Unspecified |
| I64*** | Stroke, Not Specified As Haemorrhage Or Infarction *(see note below)* |
| I676 | Nonpyogenic Thrombosis Of Intracranial Venous System |

*** NOTE: The decision to place I64 under the Ischemic group is based on the following rationale:

*Coding guidelines direct health information management systems to assume “ischemic” if hemorrhagic stroke has been ruled out (there is no supporting documentation of a hemorrhagic stroke). Hemorrhagic strokes are fairly well documented.*

*From the Canadian Institutes of Health Information (CIHI) eQuery 44220: One valid circumstance [for coding I64] is when diagnostic imaging has not yet been performed and another is when any transfer information does not indicate the type of stroke. For the majority of cases, the coder can locate the information required to assign a code for hemorrhagic stroke or ischemic stroke from the documentation. Despite specific resources (CIHI eLearning Different Codes for Different Strokes, eQueries), I64 is still frequently assigned. In such cases, the stroke was most likely ischemic.*

The following additional selection criteria were used: (a) All first and recurrent stroke events, (b) Age at admission 20 years and older, (c) Admission to an acute care facility, (d) Valid health care number (ULI), (e) Sex recorded as male or female, and (f) all discharge dispositions.

A patient was considered to have a minor stroke if they had suffered an ischemic stroke and were discharged home directly from the emergency department.

Authors AG, JMS, and MDH had full access to the database population used to create the study population. The corresponding author had full access to all data in the study and had final responsibility for the decision to submit for publication.

**Sample Size Calculation**

Sample size calculations were performed using G*Power 3.1.9.7. Between April-2015 and March-2018, 576 patients received EVT for acute stroke (about 16 patients/month) of whom 326(56.6%) received alteplase (9.3/month), with the median onset-to-treatment time being 140 minutes (IQR.90-250).^16^ Using these numbers, if we estimate a 25% drop in the acute treatment volume during the COVID-19 pandemic, we would need 38 acute stroke patients in the Pandemic period to achieve 80% power using a Poisson regression analysis. Similarly, estimating a 25% greater delay in onset-to-treatment time (140 to 175 minutes, standard deviation 80 minutes) and assuming the size of the Pandemic sample is only 10% of the pre-pandemic sample, we would need at least 375 EVT patients in the Pre-Pandemic period (already accomplished) and 37 in the Pandemic period to achieve 80% power. We therefore planned to gather data until we had accrued at least 40 treated patients in the QuICR registry in the Pandemic period. Upon extracting data until 30-August-2020, we had already accrued 408 patients in QuICR during the pandemic, so we proceeded with our analyses.

**Data Availability Statement**

All data used for this study have been stored in a deidentified database, and the programming code is stored as a STATA do-file. Requests for access to the data or code used in this paper will be considered by the corresponding author.
